# CRISPR arrays as high-resolution markers to track microbial transmission during influenza infection

**DOI:** 10.1101/2022.08.11.22278625

**Authors:** Lingdi Zhang, Jahan Rahman, Lauren Lashua, Aubree Gordon, Angel Balmaseda, Guillermina Kuan, Richard Bonneau, Elodie Ghedin

## Abstract

**Background:** The microbial community present in the respiratory tract can be disrupted during influenza virus infection, leading to functional effects on the microbial ecology of the airways and potentially impacting transmission of bacterial pathogens. Determining the transmission of airway commensals, which can carry antibiotic resistance genes that could in turn be transferred to bacterial pathogens, is of public health interest. Metagenomic-type analyses of the microbiome provide the resolution necessary for microbial tracking and functional assessments in the airways.

**Results:** We obtained 221 respiratory samples that were collected from 54 individuals at 4 to 5 time points across 10 households, with and without influenza infection, in Managua, Nicaragua. From these samples we generated metagenomic (whole genome shotgun sequencing) and metatranscriptomic (RNA sequencing) datasets to profile microbial taxonomy and gene orthologous groups. Overall, specific bacteria and phages were differentially abundant between influenza positive households and control (no influenza infection) households, with bacterial species like *Moraxella catarrhalis* and bacteriophages like *Chivirus* significantly enriched in the influenza positive households. Some of the bacterial taxa found to be differentially abundant were active at the RNA level with genes involved in bacterial physiology differentially enriched between influenza positive and influenza negative samples, primarily for *Moraxella*. We identified and quantified CRISPR arrays detected in the metagenomic sequence reads and used these as barcodes to track bacteria transmission within and across households. We detected a clear sharing of bacteria commensals and pathobionts, such as *Rothia mulcilaginosa* and *Prevotella* bacteria, within and between households, indicating community transmission of these microbes. Antibiotic resistance genes that mapped to *Rothia* and *Prevotella* were prevalent across our samples. Due to the relatively small number of households in our study we could not determine if there was a correlation between increased bacteria transmission and influenza infection.

**Conclusion:** This study shows that microbial composition and ecological disruption during influenza infection were primarily associated with *Moraxella* in the households sampled. We demonstrated that CRISPR arrays can be used as high-resolution markers to study bacteria transmission between individuals. Although tracking of antibiotic resistance transmission would require higher resolution mapping of antibiotic resistance genes to specific bacterial genomes, we observed that individuals connected by shared bacteria had more similar antibiotic resistance gene profiles than non-connected individuals from the same households.

## Introduction

Influenza infection as a contagious respiratory illness causes significant morbidity and mortality worldwide. Bacterial co-infection during influenza infection, particularly in the elderly and immunocompromised populations, can play an important role in disease progression leading to complications and severe disease outcomes [1]. Infections with respiratory viruses can also disrupt the microbiome of the airways and potentially contribute to disease severity [2]. A number of studies have demonstrated viral disruption of the microbiota in the respiratory tract with changes in relative abundance of bacterial taxa such as *Pseudomonas, Corynebacterium*, and *Streptococcus* [3, 4]. The use of antibiotics, often prescribed for influenza patients because of secondary bacterial infections, can disrupt the microbiota and diminish the protective function of the microbiome [5], as well as contribute to the emergence of antibiotic resistant bacterial strains. We and others have shown that the respiratory tract can be a potential reservoir of antibiotic resistance genes in humans. Interestingly, the presence and expression of antibiotic resistance genes detected were often to drugs the subjects had not been taking [6, 7], indicating potential transmission between individuals. The transmission of opportunistic pathogens in the respiratory tract, such as *Streptococcus pneumonia*, is known to be associated with respiratory tract viral infection and younger age of the infected subject [8, 9]. For bacteria to transmit to a new host, the invading bacteria need to interact with the residing microbes and establish colonization [8, 10], more likely to occur when the microbiome in the new host is disrupted.

Despite these observations for influenza and other viral pathogens [2-4], the dynamics of the respiratory tract microbiome and the disruption of its overall microbial ecology in human seasonal influenza infection remains to be characterized. For example, very little has been reported on the modulation of phages in the respiratory tract during acute respiratory virus infection [11]. Phages, an important entity in the human microbiome, were found to shape and co-evolve with their bacterial hosts impacting bacterial growth, metabolic activities, virulence, and antibiotic resistance [12]. The compositional shift of phages and the microbiome were found to be associated with human disease, such as inflammatory bowel disease [13]. Interacting with both their bacterial hosts and eukaryotic cells, phages can potentially provide another layer of information to the characterization of the microbiome in response to influenza infection. In this study, we used CRISPR spacers to study the interactions between bacteria and phages. CRISPR functions as the bacterial immune system to defend against virus infection by integrating a 20-70 bp viral spacer into the CRISPR locus when the bacteria are first exposed to the virus. Bacteria that have the integrated sequences are then able to defend themselves against viruses that match those spacer sequences [14]. We hypothesized that this unique history record of a bacteria’s encounter with a phage could be used to profile the dynamics of the microbial ecology within the respiratory tract.

A goal of our study was to profile the disruption of the microbiome with influenza infection and to determine, from metagenomic analyses of upper respiratory tract samples, whether we could quantify transmission of commensal bacteria and antibiotic resistance genes. Currently, most studies on bacteria transmission focus on one bacteria species and use single nucleotide polymorphisms (SNPs) in marker genes [15] or whole bacterial genomes [16, 17]. If using metagenomics data, this would require very deep sequencing depth and could only sufficiently profile SNPs from the most abundant bacterial genomes. Instead, we leveraged the unique nature of bacterial CRISPR arrays as markers to track bacteria transmission of different bacteria species. Viral spacers are constantly acquired by the bacteria and integrated at the end of CRISPR arrays, proximal to the leader sequence [14]. Although the spacer sequences that the bacteria acquire from a specific virus are not entirely random, as bias in spacer sequence distribution has been observed [18, 19], the possible number of unique spacer sequences bacteria can acquire from a virus infection is large. Given the dynamics of the CRISPR arrays, the probability that individuals share the exact same sequences due to independent spacer integration events is negligible. We demonstrate that CRISPR arrays can indeed be used to study bacteria transmission with better resolution than SNP-type analyses, especially when the CRISPR array is large.

## Results

### Study cohort and sample collection

We obtained 221 respiratory samples (pooled nasal and throat swabs) that were collected from 54 individuals participating in the Household Influenza Transmission Study (HITS) in Managua, Nicaragua. In total, 10 households with 4-8 members in each household participated in the study, and samples were collected at 4 to 5 time points for each individual, at 2-4 day intervals. Sample collection was independent of influenza infection, thus some of the samples were collected at time points when the individual was not infected or had recovered (**Table S1**). The households were assigned to high, low, or no influenza virus (control) infection groups based on the number of individuals per household who tested positive for influenza. High infection households had all or 2/3 of the household members testing positive at some point over the serial sampling (5-8 household members), while the low infection households had less than a third of household members testing positive for influenza at any time point (2-3 members). The ‘no flu’ households represent uninfected controls (**Table S1**). We did not sample all the household members from the low influenza and control households. Influenza infection was diagnosed by rtPCR and the infections were all due to influenza A virus subtype H3N2. Total RNA and DNA were extracted from each sample and processed for RNAseq (metatranscriptomics) and whole genome shotgun (metagenomics), respectively, for an in-depth microbiome analysis of the upper respiratory tract across household members. Of the 221 samples, we obtained 167 metagenomics and 178 metatranscriptomics libraries; 135 samples had both metagenomics and metatranscriptomics data and we focused on these samples for a subset of the analyses.

### Impact of influenza infection on microbial composition in the upper airways

To profile the microbial composition in subjects with and without influenza across the households studied, we did a taxonomic classification on the metagenomics and metatranscriptomics reads post filtering (human reads were removed from both datasets and rRNA reads were removed from the metatranscriptomics dataset). We detected bacteria, phages, and eukaryotic viruses across the samples (Human viruses are shown in **Tables S2 and S3**) by using kraken2, which is based on exact k-mer matches to reference genomes [20]. Of the human viruses detected, beta herpesvirus was the most prevalent across samples. There was no correlation between viruses detected and the influenza infection status of the individuals, except for influenza A virus sequence reads, which were, as expected, enriched in the flu positive samples, validating the quality of the metatranscriptomics datasets (Fisher’s Exact test and FDR <=0.05).

To analyze the microbial community across the household groups and influenza infection status, we compared the relative abundance of bacteria and viruses for different comparisons; the household subjects and samples are summarized in **Table 1** and **Table 2**. Children were enriched in the flu infection group as they were the index cases and the first household members tested to have influenza infection. Pooling samples from different timepoints for each individual, we identified significant differences in bacteria diversity between household groups at both DNA and RNA levels (PERMANOVA [21] p value=0.0005 and 0.043 for metagenomes and metatranscriptomes, respectively; **Fig. 1a** and **1b**). We applied differential expression analysis (DESeq2 [22]) to identify specific bacterial taxa that drove the differences in the beta diversity. *Dolosigranulum* was significantly enriched in the metagenomes of the control households compared to the low infection households but enriched in the metatranscriptomes of the high flu infection households compared to the low infection households (**Fig. 1c** and **1d**). *Moraxella* was enriched in the high flu infection households for both metagenomes and metatranscriptomes (**Fig. 1c** and **1d**).

**Table 1.** Summary table for individuals across the households.

| Flu infection |  |  |  |  |
| --- | --- | --- | --- | --- |
| Number of Households | High Infection<br>N=4 | Low Infection<br>N=4 | No infection<br>N=2 | p values |
| Number of individuals | 23 | 23 | 6 |  |
| Age Median (sd) | 8 (6.5) | 13 (9.5) | 7 (8.3) | 0.0981 |
| Gender |  |  |  | 1 |
| Female (%) | 16 (70) | 17 (70) | 4 (70) |  |
| Male (%) | 7 (30) | 6 (30) | 2 (30) |  |

The p values were calculated by using ANOVA and Fisher's Exact tests.

**Table 2.** Summary table for flu infection and no infection samples.

| Flu infection |  |  |  |
| --- | --- | --- | --- |
|  | Infection<br>N=42 | No infection<br>N=93 | p values |
| Age Median (sd) | 6 (5.4) | 14 (8.2) | 4.47E-07 |
| Gender |  |  | 0.02051 |
| Female (%) | 15 (47) | 74 (80) |  |
| Male (%) | 17 (53) | 19 (20) |  |

The p values were calculated using ANOVA and Fisher's Exact tests

**Figure 1.**
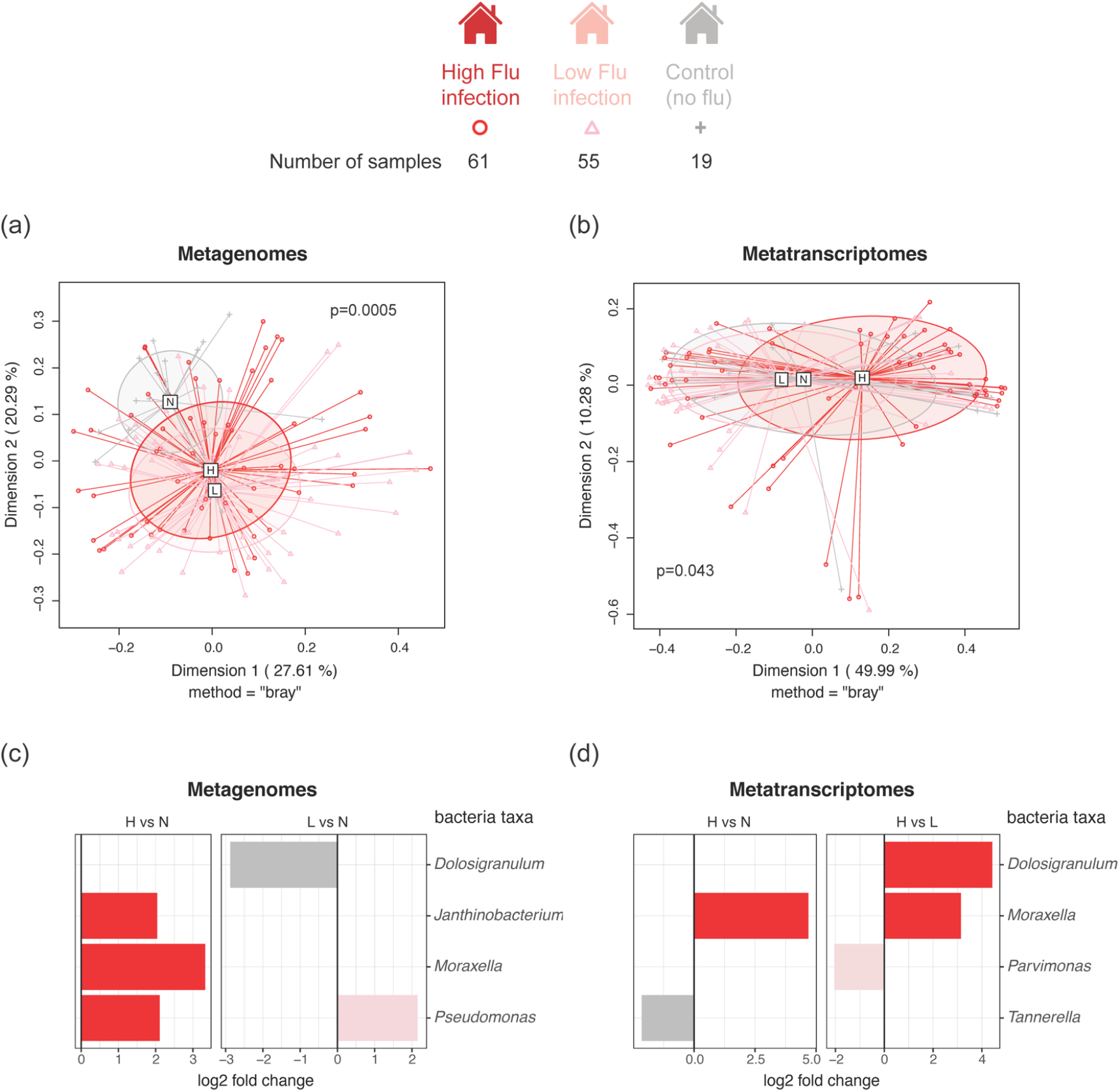
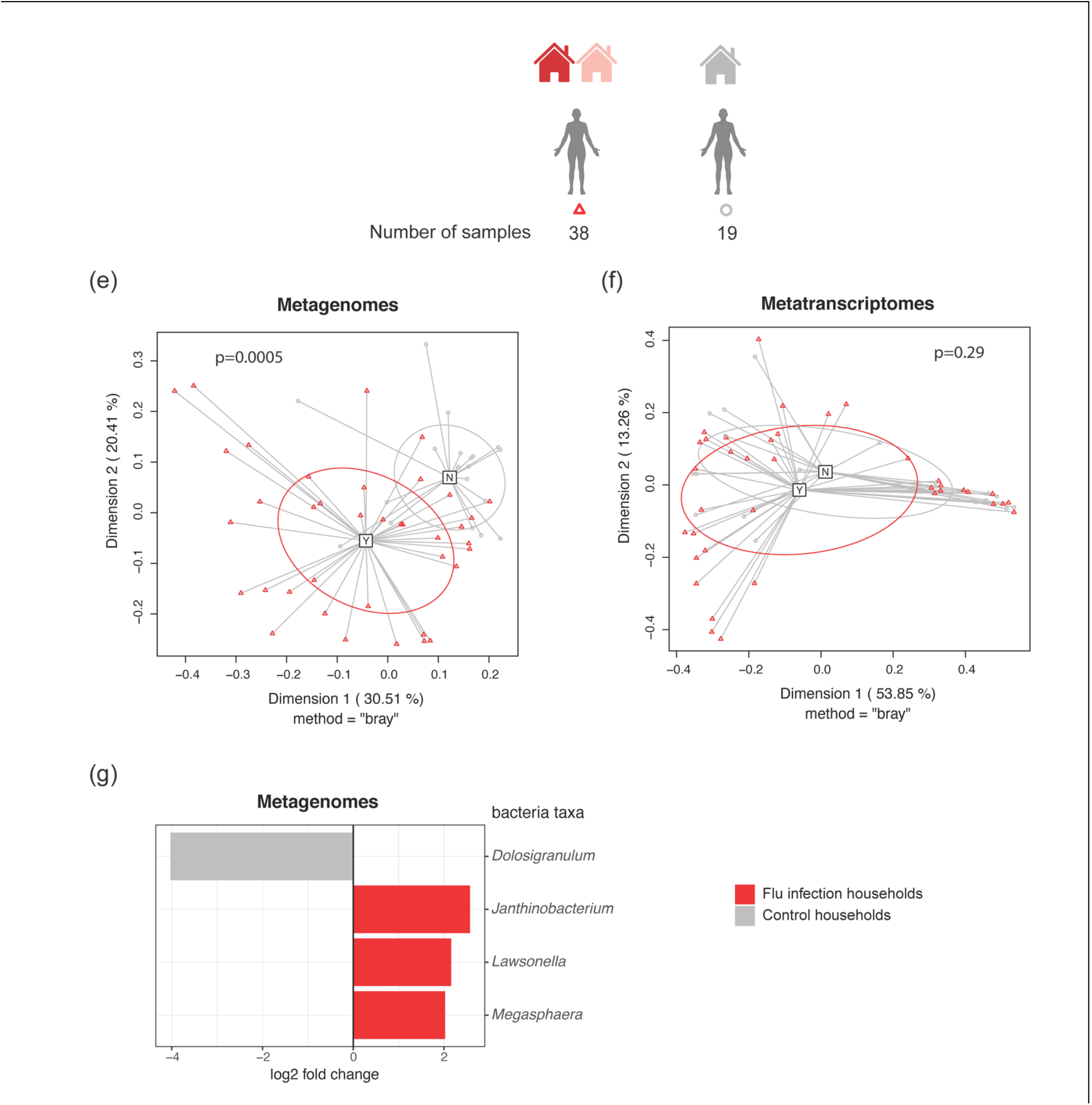
Differential abundance of bacteria and phages between households and samples. **(a)** PCA plot of β diversity of the microbial composition from metagenomics datasets for different influenza infection households. Red indicates the high flu infection households, pink indicate low flu infection households, and grey indicates control households. **(b)** PCA plot of β diversity of the microbial composition from metatranscriptomics datasets for different influenza infection households. Red indicates the high flu infection households, pink indicate low flu infection households, and grey indicates control households. **(c)** Differential abundance of bacteria genera between different household groups in metagenomes. Red or pink indicate higher differential abundance in the flu infection group tested while grey indicates it is higher in the control group. **(d)** Differential abundance of bacteria genera between different household groups in metatranscriptomes. Red or pink indicate higher differential abundance in the flu infection group tested while grey indicates it is higher in the control group. **(e)** PCA plot of β diversity of the microbial composition from metagenomics datasets for flu negative individuals from flu infection or control households. Red indicates the flu infection households and grey indicates control households. **(f)** PCA plot of β diversity of the microbial composition from metatranscriptomics datasets for flu negative individuals from flu infection or control households. Red indicates the flu infection households and grey indicates control households. **(g)** Differential abundance of bacterial genera between no flu infection individuals from flu infection and control households. Red indicates higher differential abundance in the flu infection households while grey indicates it is higher in the control group.

By comparing flu negative samples from individuals in the influenza infection households with the (flu negative) samples from the control households, we identified significant differences in bacterial composition of the metagenomes (p-value=0.0005) but not of the metatranscriptomes (p-value=0.29) (**Fig. 1e** and **1f)**. A few bacteria taxa were differentially enriched between the two groups, including *Dolosigranulum*, which was enriched in the control households (**Fig. 1g**).

We compared samples from individuals who did not test positive for influenza at any time point from any of the households (including the controls) and compared them to influenza positive samples. The microbial composition was significantly different (metagenomes p-value=0.015) but not the microbial expression profile (metatranscriptomes p-value=0.099, **Fig. 2a**). *Moraxella* was enriched in the flu positive samples (**Fig 2b**). Since *Moraxella* was enriched in the high infection households and the flu infection samples, indicating it correlated with influenza infection, we compared the relative abundance of the *Moraxella* species between flu negative time points (baseline) and flu positive time points from the same individuals. A few of the *Moraxella* spp, especially *Moraxella catarrhalis*, were moderately enriched in the flu positive time points (log2 fold change >1) in both metagenome and metatranscriptome datasets (**Fig. 2c**). We also identified phages, such as Salmonella phage Chi (or *Chivirus*) which was enriched in the high flu infection households compared to the ‘no flu’ infection households (FDR<=0.05, log2 fold change =2.17) while Pahexavirus was enriched in the low flu infection households compared to the ‘no flu’ infection households (FDR<=0.05, log2 fold change =2.69)

**Figure 2.**
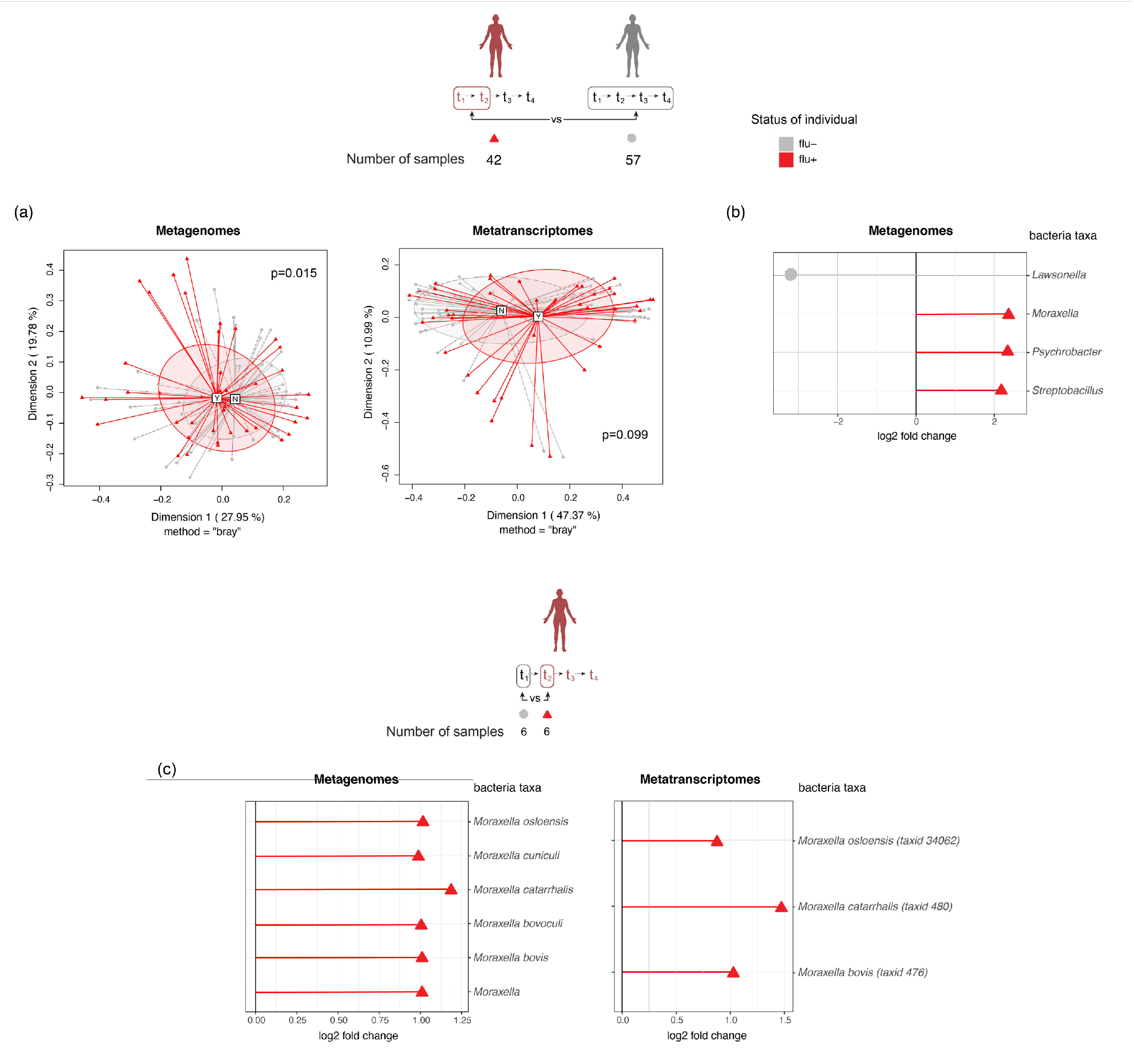
Differential abundance of bacteria and phages between influenza infection status. **(a)** PCA plot of **β** diversity of the microbial composition from flu positive and flu negative samples from metagenomics and metatranscriptomics datasets. Red indicates flu positive samples and grey indicates flu negative samples. **(b)** Bacterial genera differentially abundant in metagenomes between flu positive and flu negative samples; red indicates the taxa are enriched in flu positive samples and grey enriched in flu negative samples. **(c)** Log2 fold change of *Moraxella* species between flu positive and flu negative time points from the same individuals in metagenomes and metatranscriptomes. The red triangles indicate enrichment in the flu positive time points.

To characterize microbial functional changes during influenza infection, we profiled the relative abundance of microbial genes with FMAP, a tool that provides a functional analysis from metagenomic and metatranscriptomic sequencing data [23]. We also identified gene orthology groups (KEGG) and their relative abundance. As previously done, we partitioned the samples into influenza infection timepoints and no infection groups (only samples from individuals that did not test as flu positive were included in the ‘no infection’ group). In the metagenomics data we identified i number of microbial genes differentially abundant between flu positive and flu negative samples, including genes that allow bacteria to adapt to the environment, such as the two-component system where histidine kinase detect stimuli from the environment and trigger downstream regulatory responses [24] (**Fig. 3a**). We did not identify genes differentially expressed in the metatranscriptomics data. To identify which specific bacterial taxa contributed to the differentially abundant genes between flu positive and flu negative samples, we extracted the reads mapped to these genes and assigned taxonomic classification using Kraken2. *Moraxella* was the dominant genus contributing to the differentially abundant genes (**Fig. S1)**, with *Moraxella catarrhalis* largely contributing to these genes in the flu positive samples (**Fig. 3b**).

**Figure 3.**
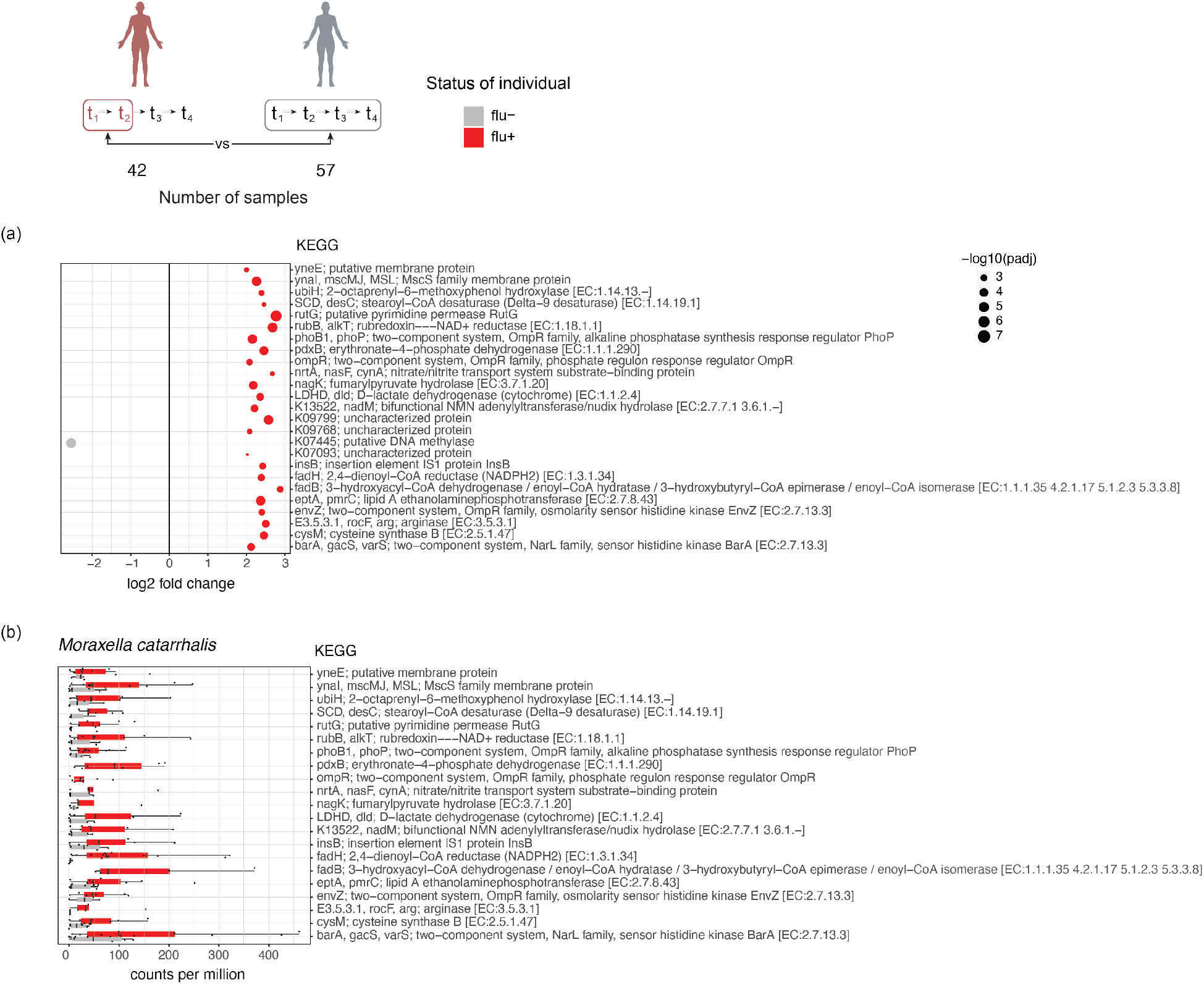
Bacteria genes and pathways altered in influenza infection. Microbial gene orthologous groups were profiled for each sample. Microbial orthologous genes differentially abundant in metagenomes between flu positive samples and flu negative samples from all households were identified using DESeq2 with a p value smaller than 0.05 and a log 2 fold change larger than 2 or smaller than -2. **(a)** The dot plot shows differentially abundant genes between flu positive and flu negative samples. The size of the dot indicates – log10 of the adjusted p values. Red indicates that the relative abundance of the gene is higher in flu positive samples and grey indicates it is higher in flu negative samples. **(b)** The reads mapped to the differentially abundant genes in (a) were extracted and their taxonomy classifications were determined through Kraken2. *Moraxella* contributes the most to these genes. The graph shows the boxplot of counts per million for the reads mapped both to *Moraxella catarrhalis* and the genes on the y axis. The samples were separated into the flu positive or flu negative groups. Red indicates flu positive samples and grey indicates flu negative samples.

### Shared CRISPR spacers to identify transmission events

We used the metagenomics datasets to study phage-bacteria interactions by focusing on the CRISPR array-integrated spacers. As phages play an important role in shaping the bacterial population and could affect host immunity, we identified phages differentially abundant between influenza positive households and control households. We profiled phage-bacteria interactions to further investigate the microbial ecological changes associated with influenza infection and to identify bacterial host species for the enriched phages found. The spacers in the CRISPR arrays, originating from viruses and integrated into the bacterial genomes, were used to link the bacteria and virus. The spacer sequences were mapped back to the viral and bacterial contigs, leading to the identification of several bacteria and phages that were linked by the shared spacers. Phages were connected to bacterial hosts both in and outside their designated host range (**Fig. S2)**. Some interactions between bacteria and phages were shared between individuals and others were unique to an individual (**Fig. S2**). A few bacterial commensals and pathobionts in the respiratory tract, such as *Prevotella* and *Rothia mucilliginosa*, were found to be infected by many phages. No specific phage-bacteria interactions appeared to be associated with influenza infection (determined by Fisher’s Exact test). This indicates either that influenza infection does not disrupt the phage-bacteria interaction dynamics in the respiratory tract or the interactions we profiled happened before influenza infection.

One important question when considering the disruption of the respiratory microbiome in a respiratory viral infection is whether certain bacteria with pathogenic potential are more likely to be transmitted. Since many commensals and pathobionts are natural members of the respiratory community [25, 26], determining the dynamics of respiratory commensal bacteria transmission within households is challenging. We used the CRISPR spacers and arrays to track bacteria transmission. Bacteria with the same set of spacers in the same order are likely to be related, indicating a potential transmission event if the same CRISPR arrays are found in two individuals. We first identified spacer sequences from the metagenomics reads using MetaCRAST. We pooled the spacers across all the samples and identified spacers shared between samples based on 90% sequence similarity. We then determined the proportion of spacers that were shared between any two samples. Samples from the same individual collected at different time points shared more spacers than samples from different individuals (**Fig. 4a**). Although spacer content is dynamic over time within individuals because of continuous interactions between phages and their bacterial hosts, the spacers were not significantly different over the sampling period. We also found that the proportion of shared spacers was higher when comparing samples from individuals living in the same household and individuals from different households, which is what we would expect with transmission (**Fig. 4a**). To further compare spacers identified from individuals within and across households, we pooled the serial samples for each subject and redid the analysis in subject-to-subject comparisons. Individuals from the same households have a higher proportion of shared spacers than individuals from different households (**Fig. 4b**), indicating more shared bacteria. As the number of subject-to-subject comparisons is not balanced for within and between households, we removed comparisons between individuals with lower than 2% shared spacers (**Fig. 4b**), leading to an equal number of comparisons within and between households, helping us to weigh comparisons between individuals from the same and different households equally and removing noise. A connection network was then generated based on the percent of shared spacers between individuals (**Fig. 4c**) where the nodes are the individuals and the edges are weighted by the value of the percent of shared spacers. We also detected subnetworks within the network using the shared spacer data between individuals **(Fig. 4c**). The correlation between the partition of the nodes to the subnetworks and the household metadata was 0.79, indicating the individuals within the same households are more tightly connected based on their percent of shared spacers.

**Figure 4.**
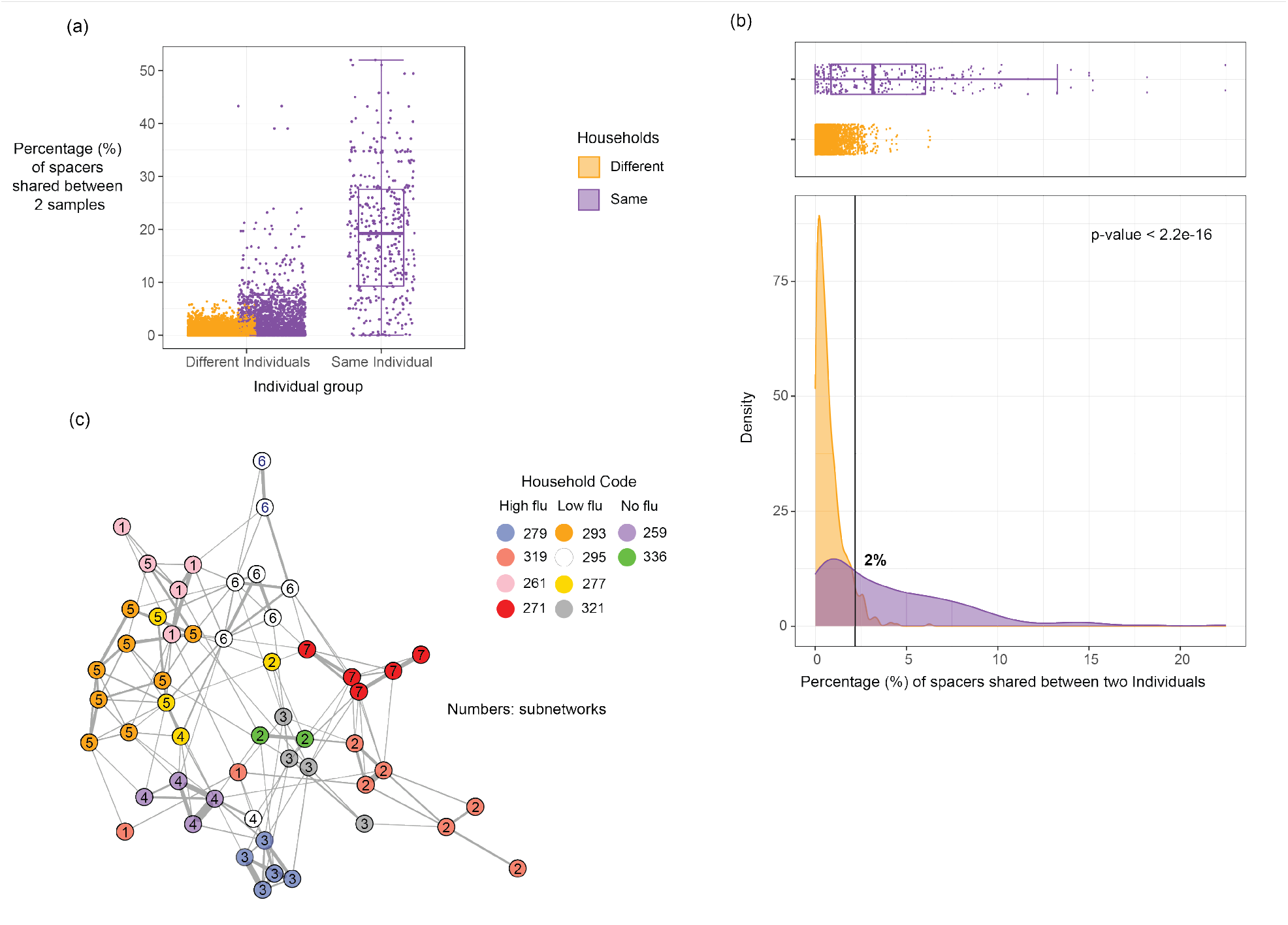
CRISPR spacers shared between samples and individuals. The percent of shared spacers between samples or individuals were compared and used to construct the connection network between individuals. **(a)** Boxplot indicating percent of spacers shared between samples from the same individuals, different individuals in the same households, and individuals from different households. The colors indicate whether the samples are from the same households (purple) or different households (orange). **(b)** Density plot and boxplot for percent of spacers shared at the individual level within and between households. The black line on the density plot indicates the cut-off where there is the same number of comparisons within and between households. **(c)** The connection network was generated based on the percent of shared spacers between individuals for the data above the cut-off in (b). The nodes represent individuals, and the edges represent percent of shared spacers. Same color nodes indicate individuals come from the same household and the numbers on the nodes represent the subnetwork they were partitioned into.

To determine which bacteria taxa were shared between individuals, we then investigated the genomic sequence where the shared CRISPR arrays were located. To do so we assembled the metagenomics reads into contigs from samples from the same individual and mapped the spacers back to the contigs to identify CRISPR arrays and the order of the spacers. We used a dynamic programming local alignment method to find the best alignments between any two CRISPR arrays that come from different individuals. We only focused on the CRISPR array alignments with more than 5 spacers. We analyzed the 2-dimensional density distribution of the CRISPR array alignments for their alignment similarity and alignment length and compared the density distributions for the alignments with CRISPR arrays from individuals from the same or different households (**Fig. S4**). Using this approach, we found that density distributions are different between the two groups we compared (**Fig. S4)** and statistically significant by using Kolmogorov-Smirnov test (p-value < 0.05) (**Fig. 5a**). We identified a region on the density plot (the upright solid purple region) that was enriched with alignments from the same households, and the alignments in that region have higher alignment similarity and alignment length (**Fig. 5a**), giving us better confidence that the CRISPR arrays in the alignments in this region are likely to be transmitted between individuals. We filtered the alignments with a similarity greater than 0.9 and an alignment length greater than 15. We extracted the contigs containing the CRISPR arrays from these alignments and aligned the contigs to the NCBI nt database to get taxonomic assignments. We identified contigs assigned to bacterial commensals and pathobionts that were found to be shared within and across households, such as *Prevotella, Fusobacterium* and *Rothia* (**Fig. 5b**). Although rare, some bacteria were shared only within households, such as *Streptococcus* and *Veillonella parvula*.

**Figure 5.**
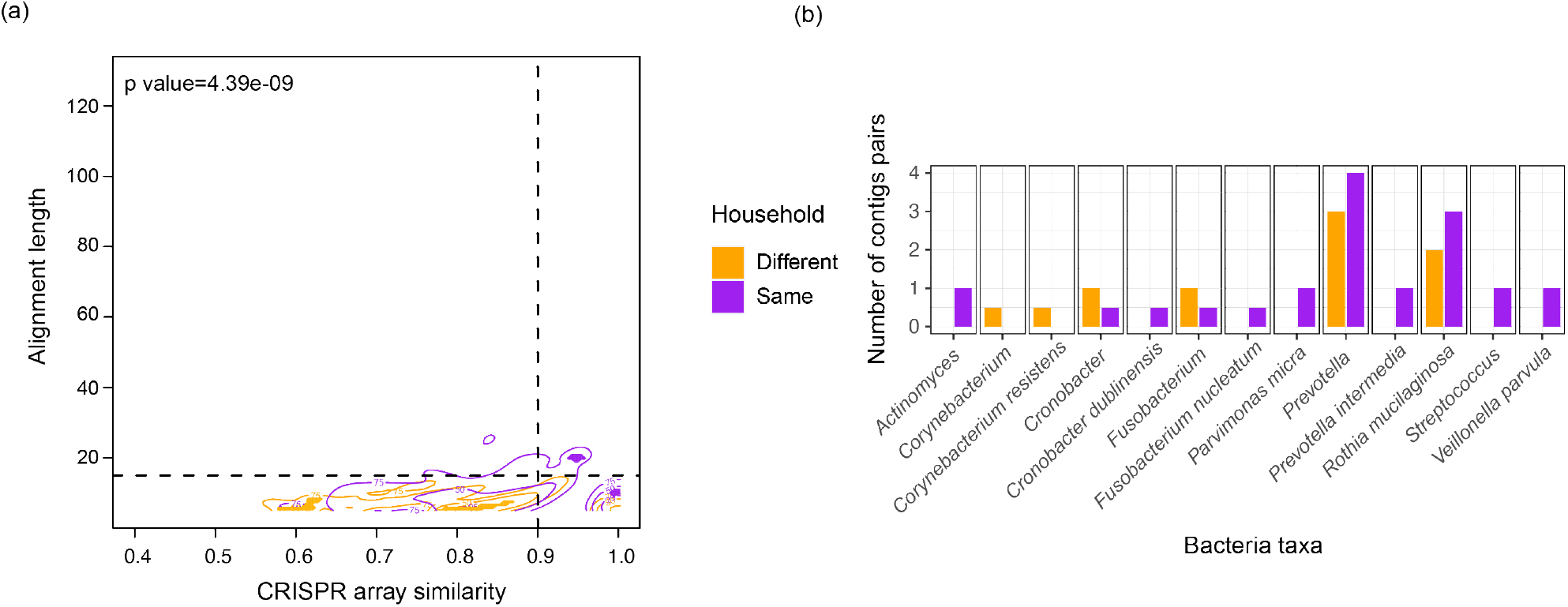
CRISPR array alignment distribution within and between households and bacteria taxa shared between individuals. **(a)** Contour plots for the 2D density distribution of CRISPR array alignments on alignment similarity and alignment length. The density distribution for the CRISPR array alignments with CRISPR arrays from individuals from the same households or different households were colored in purple and orange, respectively. The numbers on the contour plot indicate the regions have 25%, 50% and 75% of the data. The solid color regions on the plot indicate the density of the data in these regions were significantly different between the two groups. The purple region has higher data density in the same household group and the orange region has higher data density in the different household group. **(b)** The contigs contain the CRISPR arrays from the alignments with a similarity greater than 0.9 and more than 15 spacers mapped to the NCBI nt database. The graph shows the barplot of the number of individual pairs that share the bacteria with the color indicating they are from the same households or different households.

### Correlation between transmitted bacteria and shared antibiotic resistance genes

Using the spacer profile information, we first tested if there was overrepresentation of shared bacteria within flu infection households as compared to the control households. To do this we removed individual pairs that shared less than 6% of the spacers (**Fig. S5a**), which eliminated all pairs across households and helped focus on within household transmission. We analyzed pairs of individuals within the same households who shared >6% of their spacers (**Fig. S5b**) and compared across the flu infection groups the proportion of individuals in each household who passed this filter. Since we only had 10 households and there were variations in each flu infection group for the data we compared (**Fig. S5c**), we were not able to determine whether there was a correlation between bacteria transmission and flu infection levels.

We constructed a network to analyze the individuals that shared CRISPR arrays (**Fig. 6**). We looked at antibiotic use history (**Fig. S6**), influenza infection status, and age (we used 18 years old as the cut off for adults). We found that individuals connected in the network could be flu positive or flu negative, with no over representation in either group (**Fig. 6**). To detect the presence of antibiotic resistance genes, we assembled contigs across individuals (different time points from the same individual were pooled together) for both metagenomics and metatranscriptomics datasets. Antibiotic resistance genes were identified by aligning the predicted open reading frames on the assembled contigs to the CARD database [27]. The sequences predicted as antibiotic resistance genes were also filtered such that the reference genes in the database were at least 80% covered over their sequence length. By mapping the reads from the samples back to the antibiotic resistance genes detected in each subject, we quantified the antibiotic resistance genes by relative abundance. We aggregated the relative abundance values to the level of gene families that summarized the gene variants. We applied DESeq2 on the samples for both metagenomics and metatranscriptomics datasets to identify antibiotic resistance genes that were enriched or differentially expressed during flu infection. TEM and CfxA4 beta lactamase, as well as sulfonamide resistant genes (sul) were differentially abundant in the metagenomics datasets (FDR p-value < 0.05) (**Fig. S7**) but with a small log2 fold change, indicating a marginal yet statistically-significant difference. We did not detect any genes differentially expressed in the metatranscriptomics dataset. This indicates that influenza infection does not appear to be associated with increased presence of antibiotic resistance.

**Figure 6.**
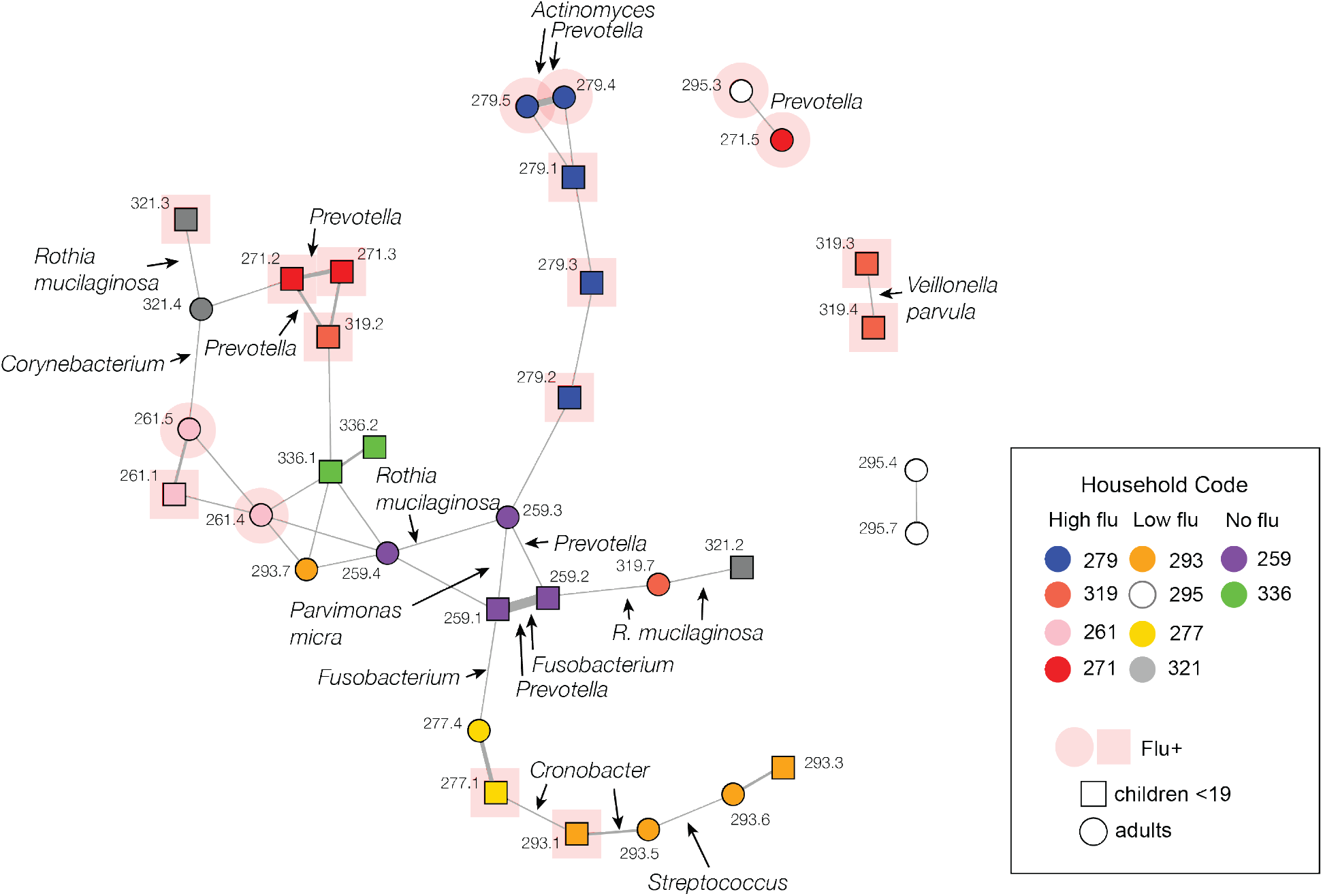
Network connecting individuals with shared CRISPR arrays. Individuals sharing CRISPR arrays were linked with the subject IDs (which are anonymized) shown next to the nodes. The color of the nodes indicates household information. The bacteria taxa annotation of the contigs containing CRISPR arrays shared between the individuals are shown next to the edges. Circle and box indicate whether the subjects are children or adults; pink color is to highlight flu positive individuals.

We used the antibiotic resistance gene profiles to calculate the fraction of individuals with a specific antibiotic resistance gene in each household, compared across the high influenza infection, low influenza infection and the control households for both metagenomics and metatranscriptomics datasets. We did not find enrichment of any genes in any group, indicating there was no differential patterns in the presence of antibiotic resistance genes among households (**Fig. S8**). Some antibiotic resistance genes were annotated with bacteria origins using the contigs to which the genes belonged (**Table S4**). We compared the antibiotic resistance gene profiles between individuals from the same households and grouped individuals on whether they were connected by shared bacteria or not. We observed that individuals connected by shared bacteria had more similar antibiotic resistance gene profiles than with other individuals from the same households, demonstrated by a smaller dissimilarity index (the two distributions were tested to be different with a p-value of 0.003) (**Fig. S9**). This implies that there might be transmission of antibiotic resistance genes together with the transmission of bacteria.

## Discussion

The respiratory tract microbiome, because of its function in health [25], should play an important role during respiratory tract infections. Here, we generated metagenomic and metatranscriptomic datasets from nasal and throat swabs to profile bacteria taxa and investigate ecological and functional aspects of the microbiome in the upper airways, as well as establish whether transmission of bacteria could be tracked using the metagenomics data.

We observe that influenza positive households were significantly different in microbial composition from the control (flu negative) households with a few bacteria and phages differentially abundant between the groups. We observed enrichment of *Moraxella* in the high flu infection households and flu positive samples. *Moraxella* has previously been found to be enriched in influenza infection samples [3], supporting our observation. *Moraxella Catarrhalis* was also enriched in the flu infection time points compared to the baseline in the same individuals. *Moraxella catarrhalis* is a common respiratory tract bacteria usually found in children that can potentially cause infections and lead to pneumonia [28]. However, one interesting observation is that the enrichment of certain bacteria that were previously thought to be associated with influenza infection, such as *Dolosigranulum* [3], were found in our study to be enriched in the control households as well, indicating that in some instances there is a household effect for the bacteria present. When comparing between influenza positive and negative time points from the same individuals, we found that the microbial diversity was not significantly different. A recent study found divergence in the respiratory tract microbiome in ferrets over 14 days post influenza infection, where the microbiome at the initial infection time points and day 14 were more like the microbiome in the uninfected ferrets [4]. In our study, although influenza viruses were not detectable after a few days, the microbiome remained similar to the infected time points, although it is possible the microbiome would have gotten back to baseline at later time points.

We identified microbial genes that were differentially enriched in flu positive and flu negative samples that are involved in bacteria physiology. This includes the two-component systems that are ubiquitous in bacteria and found to regulate virulence and antibiotic resistance in bacteria [29]. Thus, the disruption of the microbiome by influenza infection could potentially affect the balance between bacteria competition and bacteria-host interactions. *Moraxella* was found to contribute the most to these differentially abundant genes, indicating not only its relative abundance is disrupted during influenza infection but also effects on its functional potential. We also investigated whether antibiotic resistance genes were associated with influenza infection, however, we did not find a strong relationship between the relative abundance or expression of antibiotic resistance genes and influenza infection as ARGs can be detected as often in the control group.

The use of CRISPR arrays to track bacteria movement was previously done in a study investigating environmental bacteria strains on different continents [30]. However, our study is the first to use it to track bacteria transmission using clinical samples and in short-range transmissions within and between households. By using the spacers and CRISPR arrays we have shown how we could identify more transmitted bacteria within than across households, as would be expected. However, the number of shared CRISPR arrays between households also indicates that this could be an approach to potentially track transmission of bacteria on a larger scale. The individuals connected with shared bacteria include both children and adults, infected with influenza viruses and not infected. Children within households can also drive bacteria sharing as they may have closer contact with other household members. However, we do not have bacteria isolates or longer time points before influenza infection to validate bacteria transmission and to determine whether this happened during influenza infection.

There are a few limitations in this study. First, while the use of CRISPR arrays did allow the identification of shared bacteria between individuals, not all bacteria species have a CRISPR system [31], thus our analysis is restricted to a limited set of bacteria species. Also, we do not have bacteria isolates paired with the metagenomics datasets to validate the CRISPR array analysis. Second, we were limited by the number of households in the study and thus cannot draw any conclusion between bacteria sharing and influenza infection rate. The households with high or low influenza infection only indicate the members in the households were infected with influenza but we could not track specific transmission of influenza viruses among household members. Other type of evidence, such as bacteria isolates or long read sequencing would be needed to accurately link ARGs to bacteria genomes shared between individuals.

In conclusion, the analysis of the metagenome and metatranscriptome data demonstrate that the microbiome compositional and functional potentials are altered in influenza infection. In both flu positive and control individuals we saw that commensal bacteria and potential pathobionts can be readily transmitted within and across households. This implies that antibiotic resistance genes could also be transmitted. Finally, we demonstrated CRISPR arrays are a powerful tool to track bacteria transmission between individuals, offering a novel approach to leverage metagenomics datasets.

## Data Availability

The sequencing datasets supporting the conclusions of this article are available in the Sequence Read Archive (SRA). Metatranscriptome data and the metagenome data are under BioProject PRJNA713420. Scripts generating the data are available on GitHub (https://github.com/GhedinLab/Nicaragua_microbiome_flu_analysis).

## Ethical Approval and Consent to participate

This study was approved by the institutional review boards at the Nicaraguan Ministry of Health and the University of Michigan (HUM00091392). Informed consent or parental permission was obtained for all participants.

## Consent for publication

Not applicable.

## Authors’ contributions

The project was conceived by EG and RB. LZ performed sample processing, bioinformatics analyses, and writing of the manuscript. JR assisted with the development of bioinformatics pipelines. AG, AB, and GK designed and conducted the Household Influenza Transmission Study. LL assisted with the sample organization and data management. The authors read and approved the final manuscript.

## Acknowledgements

We thank the NYU Genomics Core at the Center for Genomics and Systems Biology.

## Material and Method

### Data collection

Samples were collected from individuals participating in the Household Influenza Transmission Study (HITS) in Managua, Nicaragua between July 2013 and October 2014. The HITS sample cohort included child index cases enrolled in the Nicaraguan Influenza Cohort Study and their family members who developed influenza as well as some influenza negative control households. Respiratory specimens consisted of pooled nasal and throat swabs collected from household members every 2-4 days over a 9-12 day period. Samples were shipped to the Center for Genomics and Systems Biology, New York University, and stored at - 80 °C. The HITS study was approved by the institutional review boards at the Nicaraguan Ministry of Health and the University of Michigan. Informed consent or parental permission was obtained for all participants and children aged 6 years and older provided assent.

### RNA extraction and library preparation for metatranscriptome sequencing

Total RNA was isolated from 500uL of each respiratory sample (nasal washes) with the QIAGEN RNeasy Micro Kit (QIAGEN, Hilden, Germany) according to the manufacturer’s recommendations and stored at –80°C. No mRNA enrichment or rRNA depletion steps were performed due to the limited biomass of the starting material. NEBNext Ultra II RNA Library Prep kit (New England Biolabs, Ipswich, MA) was used to generate the metatranscriptomics libraries and each library was subjected to 14-17 cycles of PCR and adaptor concentration was diluted 1:100 or 1:200 to maintain sample and adaptor ratio. Libraries were quantified by qPCR using the KAPA Library Quantification Kit (KAPA Biosystems, Wilmington, MA) on a Roche 480 LightCycler (Roche, Basel, Switzerland); their size distributions were measured on a 4200 TapeStation using a D1000 ScreenTape (Agilent Technologies, Santa Clara, CA). Libraries were diluted to 4 nM in dilution buffer (10mM Tris, pH 8.5) and combined with equimolar input into 9 sequencing pools (20-25 libraries per pool). Paired-end sequencing (2×150 bp) was performed at the Genomics Core Facility (Center for Genomics and Systems Biology, New York University) on the Illumina NextSeq 500 instrument according to the manufacturer’s instructions (Illumina, Inc., San Diego, CA) with a few libraries sequenced on the Illumina HiSeq 2500 instrument.

### DNA isolation and library preparation for metagenome sequencing

Genomic DNA was isolated from the remaining volume of each sample with the PowerSoil DNA Isolation Kit (Qiagen) and stored at –80°C. Libraries were generated using Nextera DNA Flex library prep kit (Illumina, Inc., San Diego, CA). Libraries were quantified by qPCR using the KAPA Library Quantification Kit (KAPA Biosystems, Wilmington, MA) on a Roche 480 LightCycler (Roche, Basel, Switzerland); their size distributions were measured on a 4200 TapeStation using a D1000 ScreenTape (Agilent Technologies, Santa Clara, CA). Libraries were diluted to 4 nM in dilution buffer (10mM Tris, pH 8.5) and combined with equimolar input into 9 sequencing pools (20-25 libraries per pool). Paired-end sequencing (2×150 bp) was performed at the Genomics Core Facility (Center for Genomics and Systems Biology, New York University) on the Illumina NextSeq 500 instrument according to the manufacturer’s instructions (Illumina, Inc., San Diego, CA) with a few libraries sequenced on the Illumina HiSeq 2500 instrument.

### Metagenomics and metatranscriptomics data processing

The metagenomics reads were filtered to remove adaptors and low quality reads using Trimmomatic v0.36 [32] followed by DeconSeq2 v1.32.0 [33] to remove human reads. The metatranscriptomics reads were filtered by Trimmomatic, DeconSeq2 and SortMeRNA v2.1 [34] to remove adaptors, human reads and rRNA reads, respectively. The median reads number for metagenomes was 6.8M(IQR=9.8M) and the median reads number for metatranscriptomes was 4.9M (IQR=5.6M) post filtering.

### Bacterial taxonomic assignments and differential abundant analysis

The filtered metagenomics and metatranscriptomics datasets were run through Kraken2 v2 to classify the reads to bacterial and viral taxa. Beta diversity of the metagenomics and metatranscriptomics datasets was determined using Bray Curtis distance and the global diversity between different groups was determined by PERMANOVA. The bacterial and viral taxa differentially abundant between influenza infection and no infection samples, and different household groups were identified by DESeq2 v 1.34.0 with adjusted p values <=0.05 and a log2 fold change greater than 2 or smaller than -2.

### Bacterial gene and taxa contribution

The Functional Mapping and Analysis Pipeline (FMAP [23]) was used to identify the KEGG gene orthologous groups in the metagenomics and metatranscriptomics datasets. Gene families differentially abundant or expressed between influenza positive and influenza negative samples were identified by using DESeq2 with an adjusted p value smaller than 0.05 or a log2 fold change greater than 2 or smaller than -2. The reads mapped to the KEGG gene orthologous groups that are differentially abundant between influenza positive and influenza negative groups were extracted and mapped to bacteria taxa using Kraken2.

### Bacteria and virus interaction analysis

The metagenomics bacterial reads identified by Kraken2 were pooled across different samples from the same individual and assembled into contigs using metaSPAdes v3.12.0 [35]. The spacers were mapped back to the bacterial contigs with <=1 mismatch and no secondary mapping. All filtered metagenomics reads from the same individuals across time points were assembled into contigs. VirSorter v1.1.0 [36] was used to predict the viral contigs and we focused on the contigs predicted as category 1 and category 2, which are high confidence viral contigs. The spacers were mapped back to the viral contigs using BLAST with parameters modified for short query sequences [37]. The viral and bacterial contigs were searched by BLAST against the NCBI nt database and viral Refseq database, respectively, to get taxonomy assignments. The bacterial contigs were annotated to the level of species or genus if more than 80% of the alignments on that contig were annotated as that species or genus. The viral assignments were filtered by 90% sequence similarity with the best hits (longest alignment length and highest identity). The viral and bacterial contigs linked by at least 5 spacers were identified and presented. Figure S10 provides an overview of theanalysis steps.

### CRISPR spacer analysis and network analysis

The spacers were identified from each metagenomics dataset using MetaCRAST [38]. The spacers across all the samples were clustered and spacers with sequence similarity greater than 90% using CD-HIT [39] were determined as being the same spacers across samples. The percent of shared spacers between samples was determined as the number of shared spacers between any two samples divided by the average of total spacers in the two samples. Percent of shared spacers was compared between samples from the same individuals, different individuals in the same households and different households. When comparing at the individual level, the spacers from different time points for the same individual were combined to do the analysis. The network connecting individuals based on shared spacers was generated using igraph [40] in R studio and the edge weight was the percent of shared spacers between individuals. Subnetworks were also analyzed using igraph.

### Identification of bacteria shared between individuals by using CRISPR arrays

The spacers and the order of the spacers were determined by mapping the spacers to the bacterial contigs using bowtie2 v2.2.4 [41]. A dynamic programming Smith-Waterman algorithm was used to find the best alignments between any two CRISPR arrays. We tested a few parameters for gap opening score, match score and mismatch score. Since the results were very similar for alignment length and alignment similarity (**Fig. S3**), we chose the parameters that led to the highest number of alignments between CRISPR arrays (gap score=-1, match score=3, mismatch score=-2). The CRISPR array alignments were analyzed for alignment similarity and alignment length. 2D density distribution was estimated based on the alignment similarity and alignment length using the ks package [42] **in** R studio. The densities from the same household and different households were compared, and regions on the density plot enriched with data from the same household or different households were identified.

### Antibiotic resistance gene profiling and household transmission analysis

All filtered metagenomics and metatranscriptomic reads from the same individuals across time points were assembled into contigs using metaSPAdes [35]. Open reading frames from each contig were predicted by using MetaProdigal v2.6.3 [43]. The ORFs were aligned to the CARD v3.1.0 [27] database for ARG annotation. ORFs that covered at least 80% of the ARG reference sequences were identified as present. The contigs carrying the ARGs were mapped to the NCBI nt database; the top hits, and the taxonomic information and definition for the top hits, were extracted. The metagenomic and metatranscriptomic sequence reads were mapped back to the ARGs identified in their respective datasets. The number of reads mapped back to the antibiotic resistance genes were identified and differential abundance and expression analysis were done using DESeq2.

The beta diversity of the presence/absence of the antibiotic resistance gene profiles were measured between individuals. The dissimilarities (measured by Jaccard similarity index) between same household individuals connected by shared bacteria were compared to the dissimilarities between same household individuals not connected by bacteria. The taxa origins of the antibiotic resistance genes were investigated by searching by BLAST the contigs that carry antibiotic resistance genes against the NCBI nt database; the top hits were recorded.

## Supplementary Figures

**Figure S1. Bacteria taxa mapped to genes differentially abundant between flu positive and flu negative samples**. Dotplot for the bacteria taxa mapped to genes found to be differentially abundant between flu+ and flu-samples with bacteria taxa on the x axis and genes shown on the y axis. For each gene, the color intensity of the dots shows the fraction of reads mapped to the bacteria taxa relative to total reads contributing to that gene on the y axis.

**Figure S2. Bacteria and phage interaction network**. Bacteria and viruses were linked by spacers mapped back to both bacterial and viral contigs. The graph shows the presence of interactions between specific bacteria and phages. The phages are shown on the x axis and bacteria on the y axis. The color intensity reflects the number of individuals for which the specific bacteria/phage interactions can be detected. The bacteria and phages in the same genera or found to infect bacteria in the same genera are grouped into same panels.

**Figure S3. Parameters tested for dynamic programming to align CRISPR arrays**. Different parameters were tested in a local alignment method to find the best alignments between any two CRISPR arrays. With the alignment similarity and alignment length distributions, correlations between methods and statistics are shown in the figure.

**Figure S4. Contour plot with 2D density of the CRISPR array alignments**. The 2D density distribution was estimated for the CRISPR array alignments on the alignment similarity and alignment length. The color intensity indicates the density of the data in the regions. The two panels indicate the CRISPR array alignments with CRISPR arrays from the same or different households. The y axis was adjusted to highlight the region where most of the data points are located, although there were more points over 50 on the y axis.

**Figure S5. Bacteria transmission and flu infection. (a)** Density and boxplot plot for percent of spacers shared at the individual level within and between households. The red line on the density plot indicates the cut-off where all the “between household” individual pairs were removed. **(b)** The connection network was generated based on the percent of shared spacers between individuals for the data above the cut-off in (a). The nodes represent individuals and the edges represent percent of shared spacers. Same color nodes represent individuals come from the same household. **(c)** Dotplot for percent of individuals in each household that were connected. Number of individuals in (b) in each household were divided by total number of individuals in the households and compared across flu infection groups. The x axis indicates household code and the panels show the household from high, low, or no flu infection groups.

**Figure S6. Antibiotic use history**. The antibiotics taken by the subjects within 2 years prior to influenza infection is shown in red color. White indicates that particular antibiotic was not taken by the subject, and grey indicates antibiotics use information for that individual is not available (NA).

**Figure S7. Antibiotic resistance genes differentially abundant between flu positive and flu negative samples**. Barplot for counts per million of the ARGs identified as differentially abundant between flu positive and flu negative samples. The color indicates the influenza positive (flu+) or influenza negative (flu-) groups. The panel titles indicate the mechanism used by the genes to confer antibiotic resistance. The log2 fold change of ARGs counts for the identified antibiotic resistance genes are also shown with barplot.

**Figure S8. Prevalence of antibiotic resistance genes in households across flu infection groups**. Presence/absence of antibiotic resistance genes was determined for each individual. The ratio of individuals that have a specific ARG in each household was calculated by dividing the number of individuals carrying the gene with the total number of individuals in the households. The graph shows a dotplot for the fraction of individuals that have ARGs across the flu infection groups for metagenomics (MG) and metatranscriptomics (MT) datasets. The color indicates the data type, red for MG and blue for MT. H, L or N panel titles correspond to high, low, and no flu infection groups, respectively. The color intensity indicates the fraction of individuals that have the ARGs in the household. The red horizontal panel titles indicate the mechanisms used by the ARGs to confer antibiotic resistance.

**Figure S9. Boxplot for ARG profiles dissimilarity between individuals from same households connected by shared bacteria, same households not connected and different households**. Each dot represents the dissimilarity in ARG profiles between two individuals. The color indicates the individuals were from the same or different households. The x axis shows whether the individuals were connected by shared bacteria when they are from the same household. The y axis shows the distance in ARG profiles between any two individuals.

**Figure S10. Overview of CRISPR array analysis steps**. CRISPR arrays were used for two purposes: (1) the spacers that comprise the CRISPR arrays on bacterial contigs and that can be mapped to viral contigs, were used to link bacteria and phages to study phage-bacteria interactions; and (2) the spacers and the CRISPR arrays were used as a barcode to study bacteria transmission within and across households.

## Supplementary Tables

**Table S1. Sample metadata**.

**Table S2. Human DNA viruses identified from the metagenomics datasets**. The human viruses were identified from Kraken analysis and the presence or absence of the viruses are shown as 1 or 0.

**Table S3. Human viruses identified from the metatranscriptomics datasets**. The human viruses were identified from Kraken analysis and the presence or absence of the viruses are shown as 1 or 0.

**Table S4. Antibiotic resistance genes and contig annotations**. The contigs containing the ARGs were annotated with the bacteria genome information.

