## Supplemental Figures for "CRISPR arrays as high-resolution markers to track microbial transmission during influenza infection"

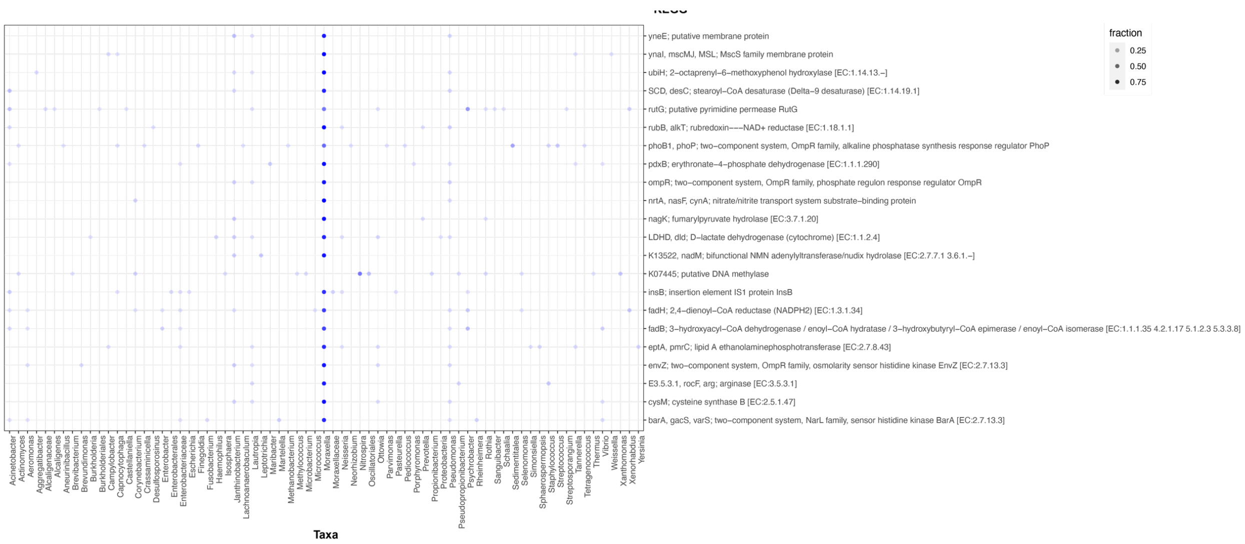


**Figure S1. Bacteria taxa mapped to genes differentially abundant between flu positive and flu negative samples**. Dotplot for the bacteria taxa mapped to genes found to be differentially abundant between flu+ and flu- samples with bacteria taxa on the x axis and genes shown on the y axis. For each gene, the color intensity of the dots shows the fraction of reads mapped to the bacteria taxa relative to total reads contributing to that gene on the y axis.

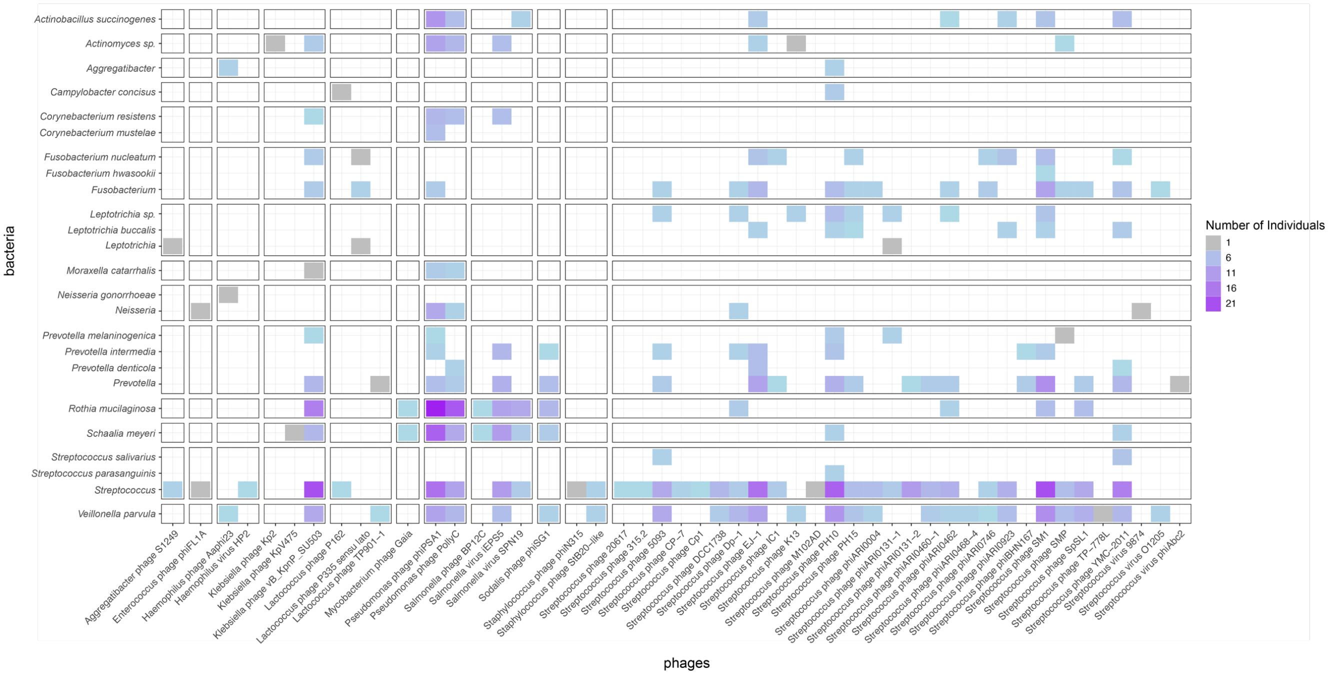


**Figure S2. Bacteria and phage interaction network.** Bacteria and viruses were linked by spacers mapped back to both bacterial and viral contigs. The graph shows the presence of interactions between specific bacteria and phages. The phages are shown on the x axis and bacteria on the y axis. The color intensity reflects the number of individuals for which the specific bacteria/phage interactions can be detected. The bacteria and phages in the same genera or found to infect bacteria in the same genera are grouped into same panels.

| | | | | | |
|---|---|---|---|---|---|
| Parameters:<br>gap=-1<br>match=1<br>mismatch=-1 | Parameters:<br>gap=-1<br>match=2<br>mismatch=-1 | Parameters:<br>gap=-1<br>match=3<br>mismatch=-2 | Parameters:<br>gap=-2<br>match=3<br>mismatch=-1 | Parameters:<br>gap=-2<br>match=2<br>mismatch=-1 | Parameters:<br>gap=-1<br>match=2<br>mismatch=-2 |
| Output:<br>Total alignments=1190<br>Alignment length stats:<br>median=9<br>similarity stats:<br>median=0.89 | Output:<br>Total alignments=1434<br>Alignment length stats:<br>median=10<br>similarity stats:<br>median=0.86 | Output:<br>Total alignments=1587<br>Alignment length stats:<br>median=9<br>similarity stats:<br>median=0.86 | Output:<br>Total alignments=1314<br>Alignment length stats:<br>median=10<br>similarity stats:<br>median=0.86 | Output:<br>Total alignments=1289<br>Alignment length stats:<br>median=9<br>similarity stats:<br>median=0.88 | Output:<br>Total alignments=1474<br>Alignment length stats:<br>median=9<br>similarity stats:<br>median=0.87 |

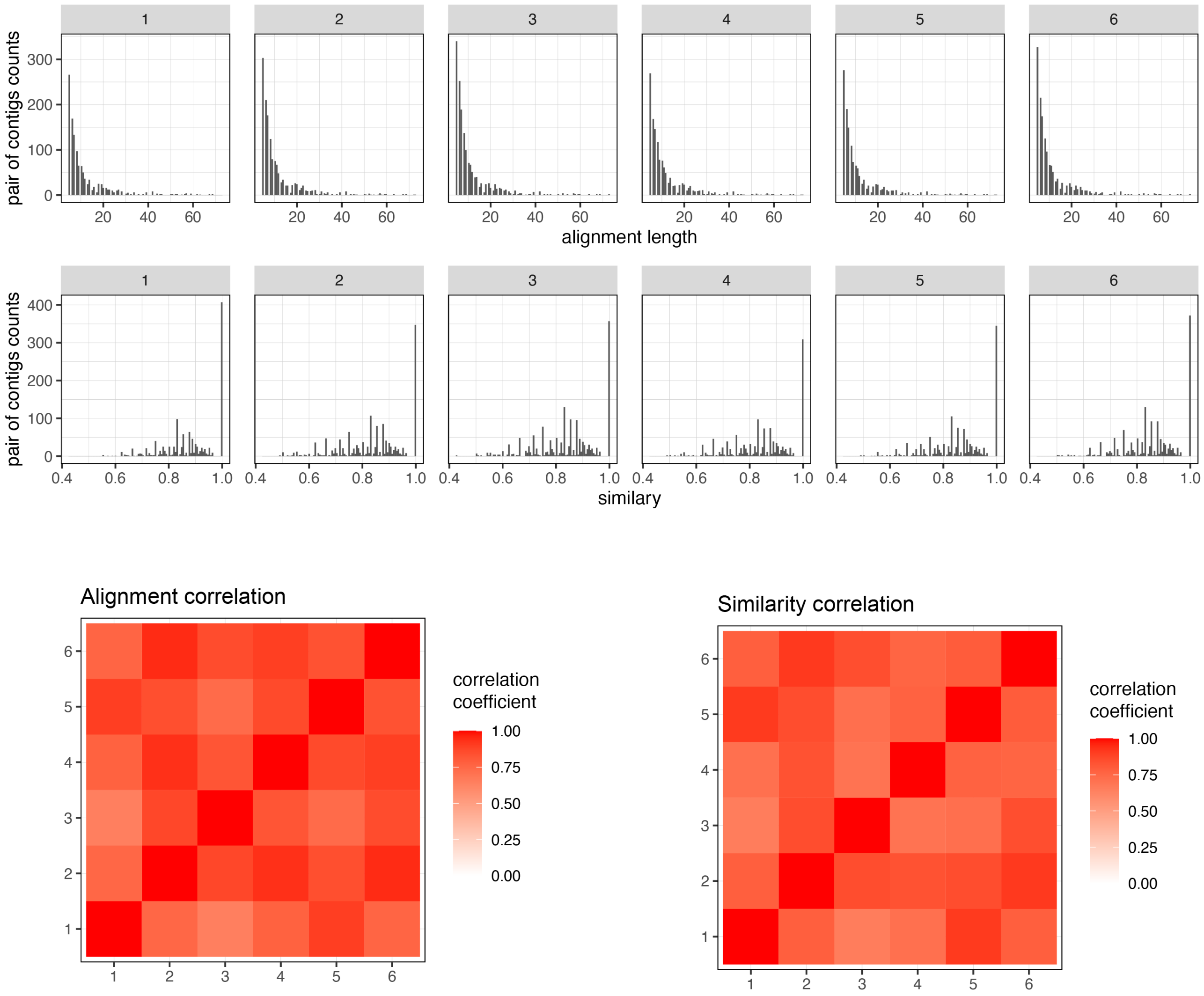


**Figure S3. Parameters tested for dynamic programming to align CRISPR arrays.** Different parameters were tested in a local alignment method to find the best alignments between any two CRISPR arrays. With the alignment similarity and alignment length distributions, correlations between methods and statistics are shown in the figure.

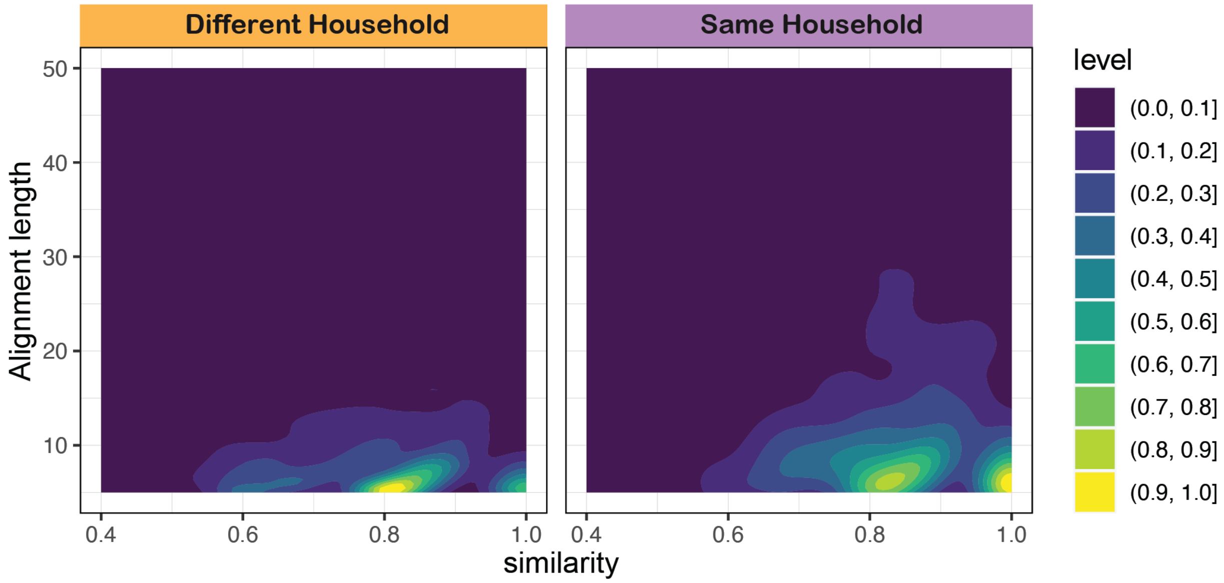


**Figure S4. Contour plot with 2D density of the CRISPR array alignments**. The 2D density distribution was estimated for the CRISPR array alignments on the alignment similarity and alignment length. The color intensity indicates the density of the data in the regions. The two panels indicate the CRISPR array alignments with CRISPR arrays from the same or different households. The y axis was adjusted to highlight the region where most of the data points are located, although there were more points over 50 on the y axis.

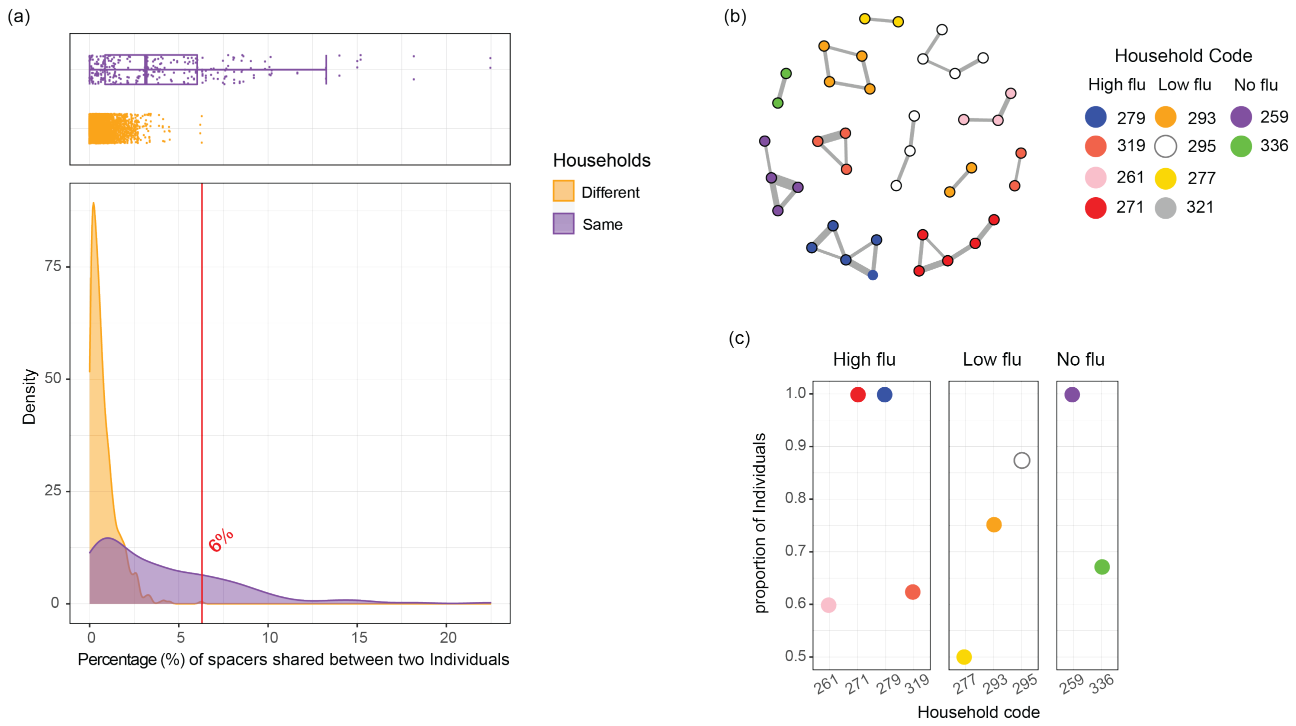


**Figure S5. Bacteria transmission and flu infection. (a)** Density and boxplot plot for percent of spacers shared at the individual level within and between households. The red line on the density plot indicates the cut-off where all the "between household" individual pairs were removed. **(b)** The connection network was generated based on the percent of shared spacers between individuals for the data above the cut-off in (a). The nodes represent individuals and the edges represent percent of shared spacers. Same color nodes represent individuals come from the same household. **(c)** Dotplot for percent of individuals in each household that were connected. Number of individuals in (b) in each household were divided by total number of individuals in the households and compared across flu infection groups. The x axis indicates household code and the panels show the household from high, low, or no flu infection groups.

**Figure S6**

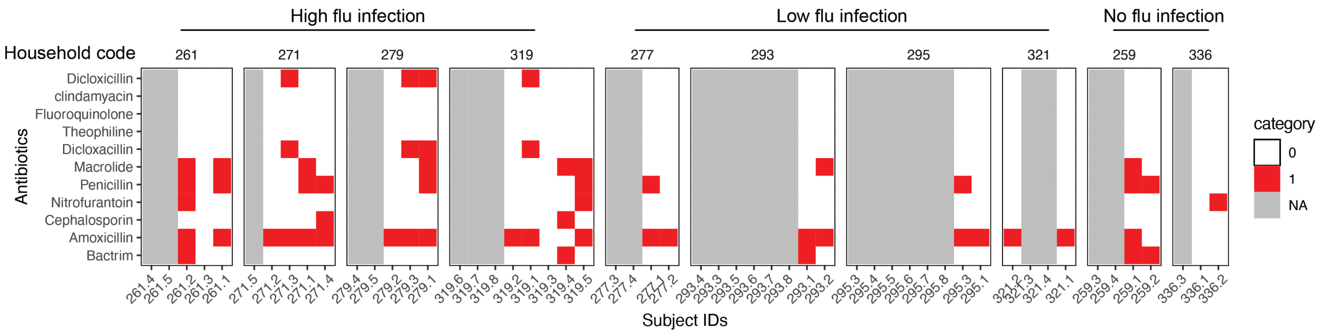


**Figure S6. Antibiotic use history**. The antibiotics taken by the subjects within 2 years prior to influenza infection is shown in red color. White indicates that particular antibiotic was not taken by the subject, and grey indicates antibiotics use information for that individual is not available (NA).

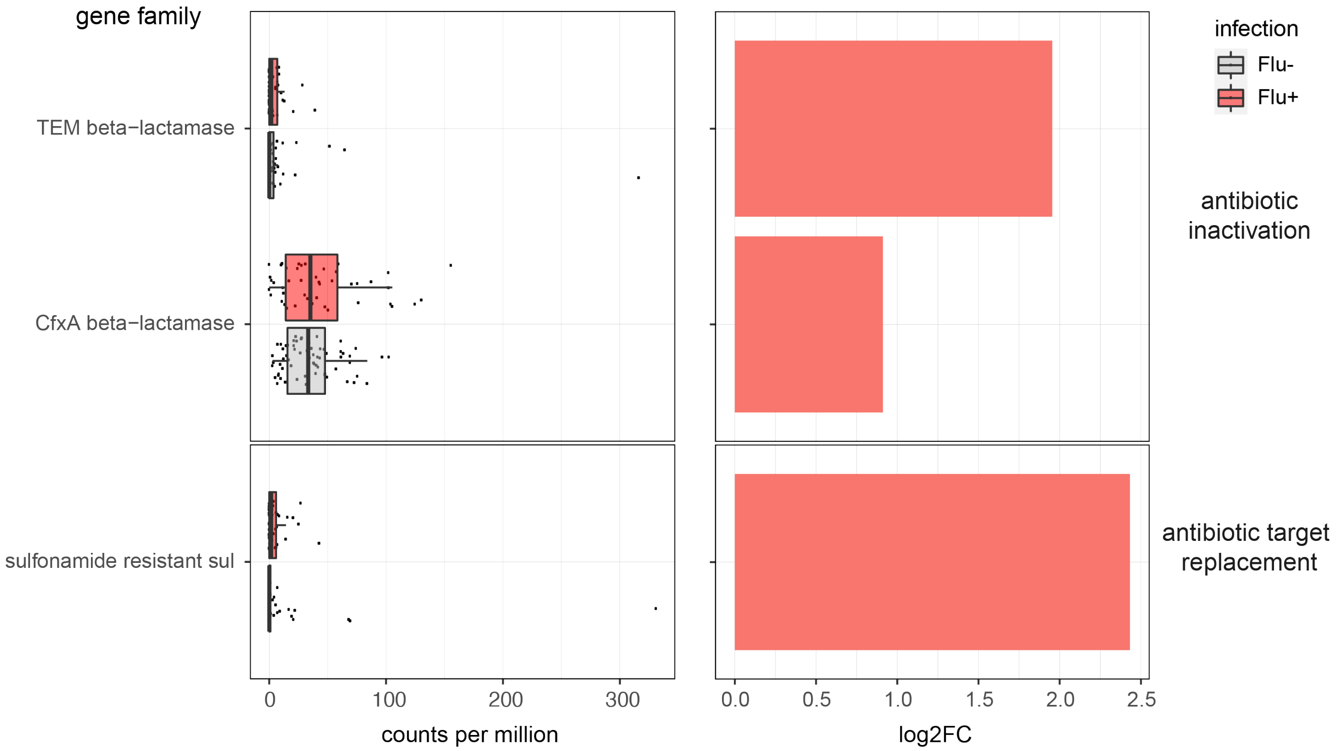


**Figure S7. Antibiotic resistance genes differentially abundant between flu positive and flu negative samples.** Barplot for counts per million of the ARGs identified as differentially abundant between flu positive and flu negative samples. The color indicates the influenza positive (flu+) or influenza negative (flu-) groups. The panel titles indicate the mechanism used by the genes to confer antibiotic resistance. The log2 fold change of ARGs counts for the identified antibiotic resistance genes are also shown with barplot.

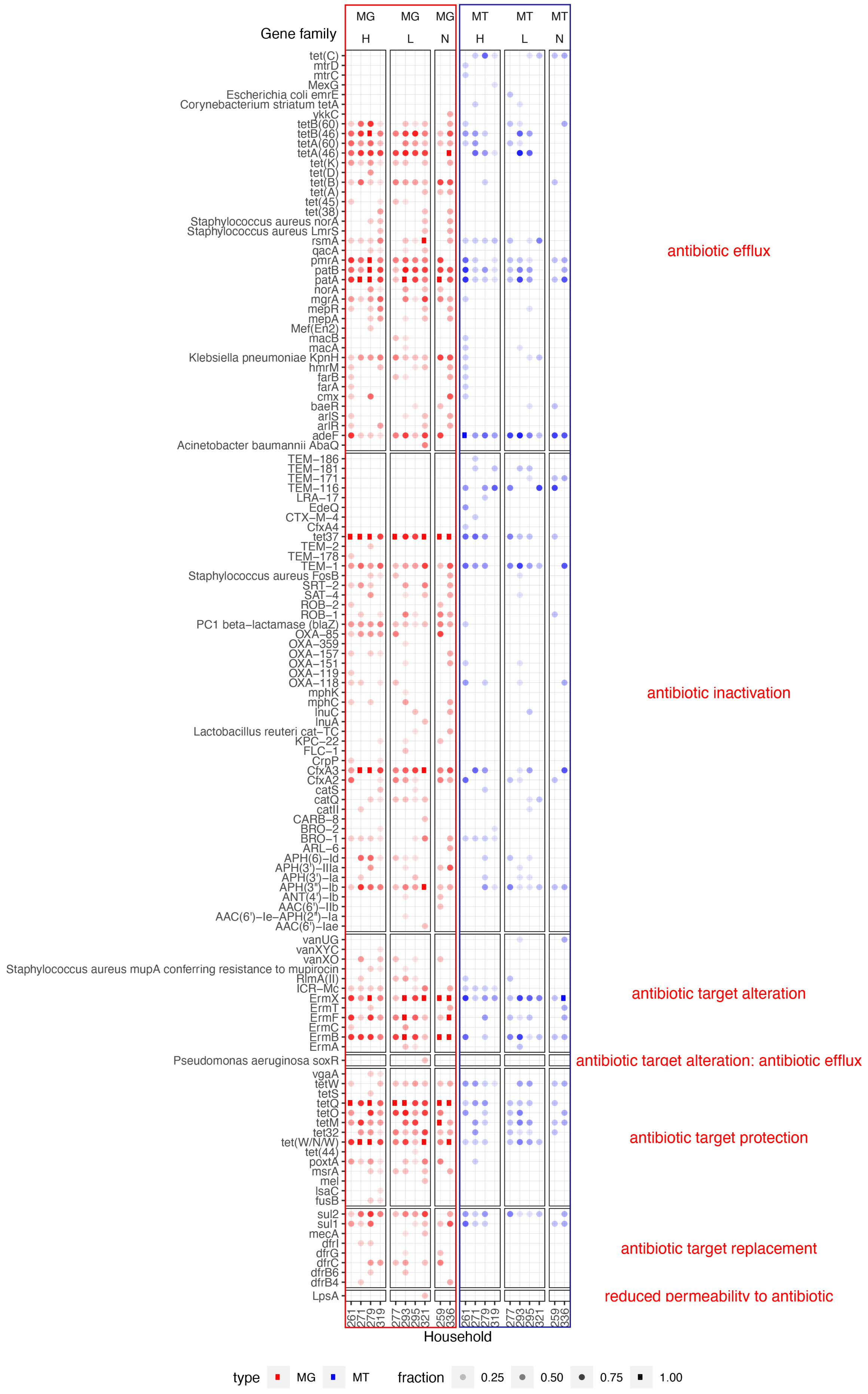
Gene family
MG H
MG L
MG N
MT H
MT L
MT N
tet(C)
mtrD
mtrC
MexG
Escherichia coli emrE
Corynebacterium striatum tetA
ykkC
tetB(60)
tetB(46)
tetA(60)
tetA(46)
tet(K)
tet(D)
tet(B)
tet(A)
tet(45)
tet(38)
Staphylococcus aureus norA
Staphylococcus aureus LmrS
rsmA
qacA
pmrA
patB
patA
norA
mgrA
mepR
mepA
Mef(En2)
macB
macA
Klebsiella pneumoniae KpnH
hmrM
farB
farA
cmx
baeR
arlS
arlR
adeF
Acinetobacter baumannii AbaQ
TEM−186
TEM−181
TEM−171
TEM−116
LRA−17
EdeQ
CTX−M−4
CfxA4
tet37
TEM−2
TEM−178
TEM−1
Staphylococcus aureus FosB
SRT−2
SAT−4
ROB−2
ROB−1
PC1 beta−lactamase (blaZ)
OXA−85
OXA−359
OXA−157
OXA−151
OXA−119
OXA−118
mphK
mphC
lnuC
lnuA
Lactobacillus reuteri cat−TC
KPC−22
FLC−1
CrpP
CfxA3
CfxA2
catS
catQ
catII
CARB−8
BRO−2
BRO−1
ARL−6
APH(6)−Id
APH(3')−IIIa
APH(3')−Ia
APH(3'')−Ib
ANT(4')−Ib
AAC(6')−IIb
AAC(6')−Ie−APH(2'')−Ia
AAC(6')−Iae
vanUG
vanXYC
vanXO
Staphylococcus aureus mupA conferring resistance to mupirocin
RlmA(II)
ICR−Mc
ErmX
ErmT
ErmF
ErmC
ErmB
ErmA
Pseudomonas aeruginosa soxR
vgaA
tetW
tetS
tetQ
tetO
tetM
tet32
tet(W/N/W)
tet(44)
poxtA
msrA
mel
lsaC
fusB
sul2
sul1
mecA
dfrI
dfrG
dfrC
dfrB6
dfrB4
LpsA
antibiotic efflux
antibiotic inactivation
antibiotic target alteration
antibiotic target alteration; antibiotic efflux
antibiotic target protection
antibiotic target replacement
reduced permeability to antibiotic
261 271 279 319 277 293 295 321 259 336 261 271 279 319 277 293 295 321 259 336
Household
type MG MT
fraction 0.25 0.50 0.75 1.00

**Figure S8. Prevalence of antibiotic resistance genes in households across flu infection groups.** Presence/absence of antibiotic resistance genes was determined for each individual. The ratio of individuals that have a specific ARG in each household was calculated by dividing the number of individuals carrying the gene with the total number of individuals in the households. The graph shows a dotplot for the fraction of individuals that have ARGs across the flu infection groups for metagenomics (MG) and metatranscriptomics (MT) datasets. The color indicates the data type, red for MG and blue for MT. H, L or N panel titles correspond to high, low, and no flu infection groups, respectively. The color intensity indicates the fraction of individuals that have the ARGs in the household. The red horizontal panel titles indicate the mechanisms used by the ARGs to confer antibiotic resistance.

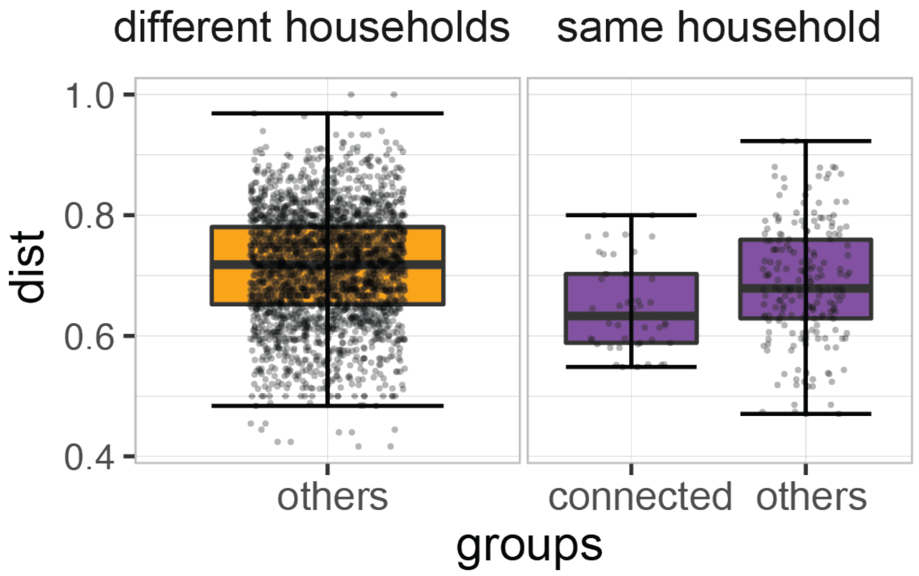


**Figure S9. Boxplot for ARG profiles dissimilarity between individuals from same households connected by shared bacteria, same households not connected and different households**. Each dot represents the dissimilarity in ARG profiles between two individuals. The color indicates the individuals were from the same or different households. The x axis shows whether the individuals were connected by shared bacteria when they are from the same household. The y axis shows the distance in ARG profiles between any two individuals.

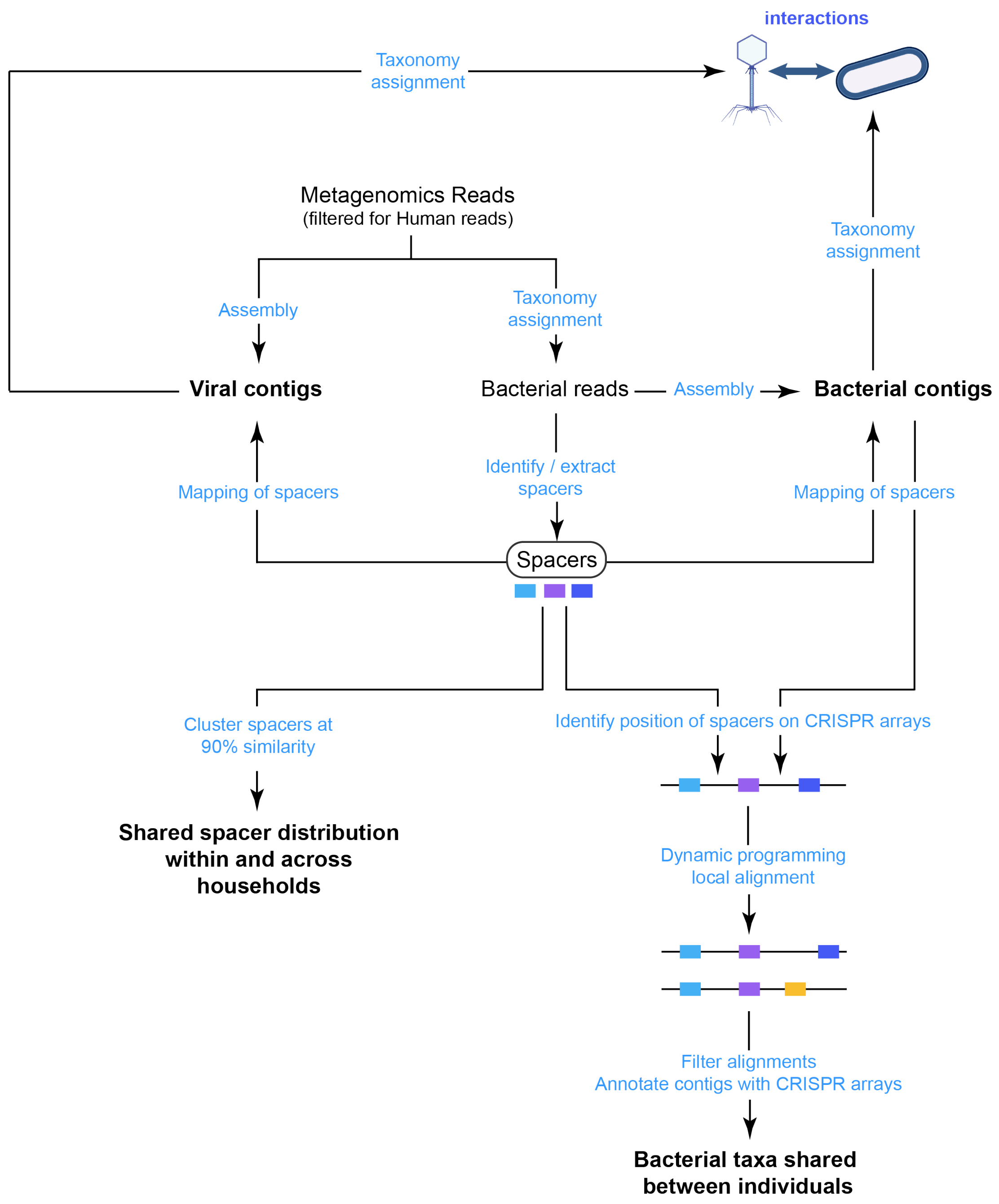


**Figure S10. Overview of CRISPR array analysis steps.** CRISPR arrays were used for two purposes: (1) the spacers that comprise the CRISPR arrays on bacterial contigs and that can be mapped to viral contigs, were used to link bacteria and phages to study phage-bacteria interactions; and (2) the spacers and the CRISPR arrays were used as a barcode to study bacteria transmission within and across households.
